# High COVID-19 vaccine coverage allows for a re-opening of European universities

**DOI:** 10.1101/2021.11.16.21266383

**Authors:** Jana Lasser, Timotheus Hell, David Garcia

## Abstract

Returning universities to full on-campus operations while the COVID-19 pandemic is ongoing has been a controversial discussion in many countries. The risk of large outbreaks in dense course settings is contrasted by the benefits of in-person teaching. Transmission risk depends on a range of parameters, such as vaccination coverage, number of contacts and adoption of non-pharmaceutical intervention measures (NPIs). Due to the generalised academic freedom in Europe, many universities are asked to autonomously decide on and implement intervention measures and regulate on-campus operations. In the context of rapidly changing vaccination coverage and parameters of the virus, universities often lack the scientific facts to base these decisions on. To address this problem, we analyse a calibrated, data-driven simulation of transmission dynamics of 10755 students and 974 faculty in a medium-sized university. We use a co-location network reconstructed from student enrolment data and calibrate transmission risk based on outbreak size distributions in other Austrian education institutions. We focus on actionable interventions that are part of the already existing decision-making process of universities to provide guidance for concrete policy decisions. Here we show that with the vaccination coverage of about 80% recently reported for students in Austria, universities can be safely reopened if they either mandate masks or reduce lecture hall occupancy to 50%. Our results indicate that relaxing NPIs within an organisation based on the vaccination coverage of its sub-population can be a way towards limited normalcy, even if nation wide vaccination coverage is not sufficient to prevent large outbreaks yet.

## Introduction

Many universities face increasing pressure to return to full on-campus operations, while COVID-19 incidence is still high. A range of simulation studies tried to assess the transmission risk and effectiveness of non-pharmaceutical intervention measures (NPIs) in the university context in the past. These studies have a number of shortcomings that limit their applicability to the decision making of universities. Only a small number of studies consider NPIs together with vaccinations (1–3) and none include the delta variant. Only a few studies base their models on empirically determined contact networks but there the transmission dynamics are either not calibrated against empirical data (4, 5) or the networks are small (4, 6). Similarly, only a handful of studies calibrate their model parameters against empirically observed outbreaks in educational settings (7–9) but these studies use simulation parameters that were determined for virus variants that are no longer dominant. In addition, existing studies focus on residential colleges and model both contacts in courses and in housing contexts. The applicability of these studies to the European higher education sector is limited, since in Europe students tend to live spread out in the university’s city. As a consequence, COVID-19 prevention policies adopted by European universities have no power to limit social contacts of students outside of courses on university premises. To our knowledge, no existing study combines an empirically determined co-location network with a rigorous calibration of model parameters and simulation scenarios that are relevant for the current decision-making processes. To remedy these shortcomings, we modelled transmission dynamics in a medium-sized European university with 10755 students and 974 faculty. We base our simulation on a co-location network reconstructed from enrolment data from the winter term 2019/20, part of which is shown in Fig. 1. At the point of writing, 82% of the students have been vaccinated (10). Based on this high vaccination coverage, we investigated whether full on-campus operation is possible, even in a situation were the delta variant is dominant.

**Fig. 1.**
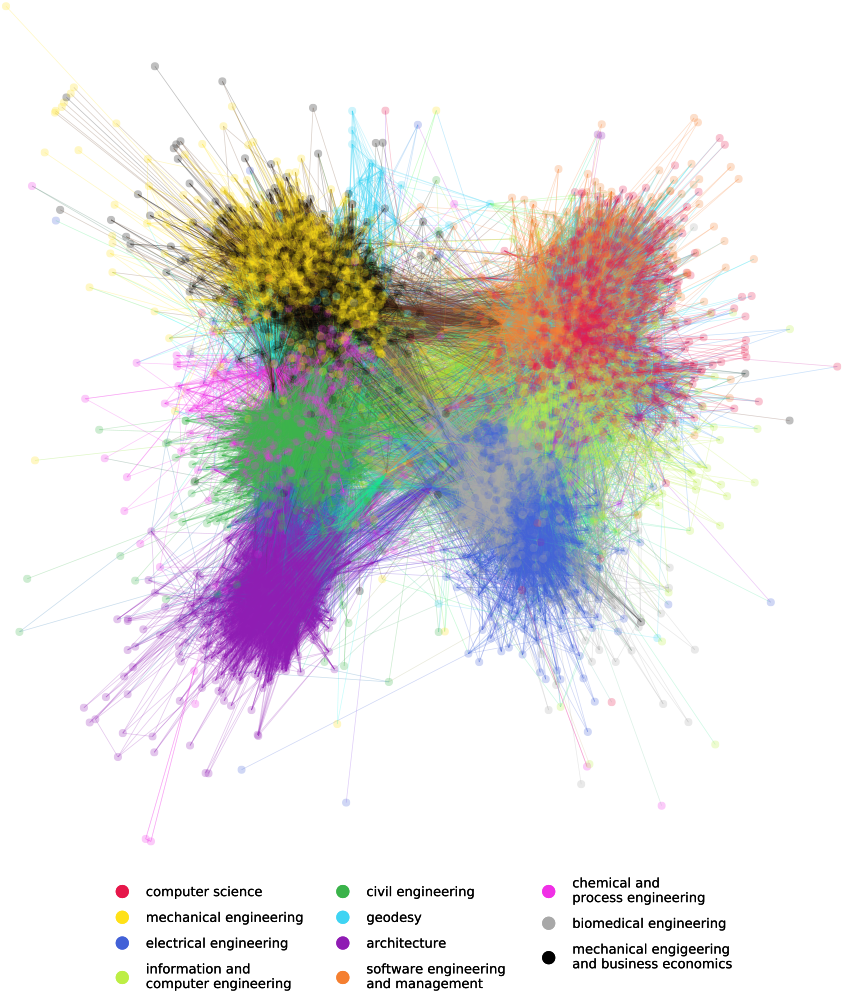
Visualisation of the co-location network of the 11 undergrad studies of TU Graz. The visualisation shows 5855 students colour-coded by their study and their connections during lectures and exams for one week in October 2019.

JL developed the research idea, implemented the simulation and wrote the initial draft of the manuscript. TH helped acquire the relevant data to construct the university contact network and defined scenarios relevant for university policy. DG provided guidance on the research questions and statistical analysis of simulation outcomes. All authors have read and edited the final manuscript.

The authors declare no competing interests.

## Results

We developed an agent-based model to simulate transmission dynamics on a co-location network determined by the interactions of students and faculty at TU Graz. We assume a vaccination coverage of 80% among students and staff (10) and a 47% vaccination efficacy against infection, as has been reported for the dominant vaccine in Austria (BNT162b2) five months after the second dose (11). We study two different lecture hall occupancy levels (50% and 100%) and masking mandates (no masks, masks for students and lectunrers). Results for these conditions are shown in Fig. 2. We show the distribution of outbreak sizes (number of infected individuals minus the source case) and the effective reproduction number *R*_eff_, i.e. the average number of agents infected by the source case. With 100% lecture hall occupancy and no masking mandate, the average outbreak size is 110 [0; 674] (95% credible interval) with *R*_eff_ = 1.05[0; 6]. The maximum observed outbreak size over 10 000 simulations is 858 and 52.5% of source cases do not infect another person. If masks are mandated, the mean outbreak size is reduced to 2.1 [0; 22] and the maximum observed outbreak size is 211, while *R*_eff_ = 0.38[0; 3] and 73% our source cases do not infect another person. On the other hand, if no mask mandate is implemented but instead lecture hall occupancy is reduced to 50%, the average outbreak size is 7.4 [0; 112] with *R*_eff_ = 0.61[0; 4]. We additionally analysed different vaccination coverage in the student population, finding that if vaccination coverage is reduced to 70%, the mean outbreak size with a general mask mandate and 100% occupancy increases to 16 [0; 267], with *R*_eff_ = 0.55[0; 4]. With 50% occupancy and no masks, average outbreak sizes reach 75 [0; 687] cases with *R*_eff_ = 0.96[0; 6]. If no students and faculty members are vaccinated, the mean outbreak size surpasses 5200 cases.

**Fig. 2.**
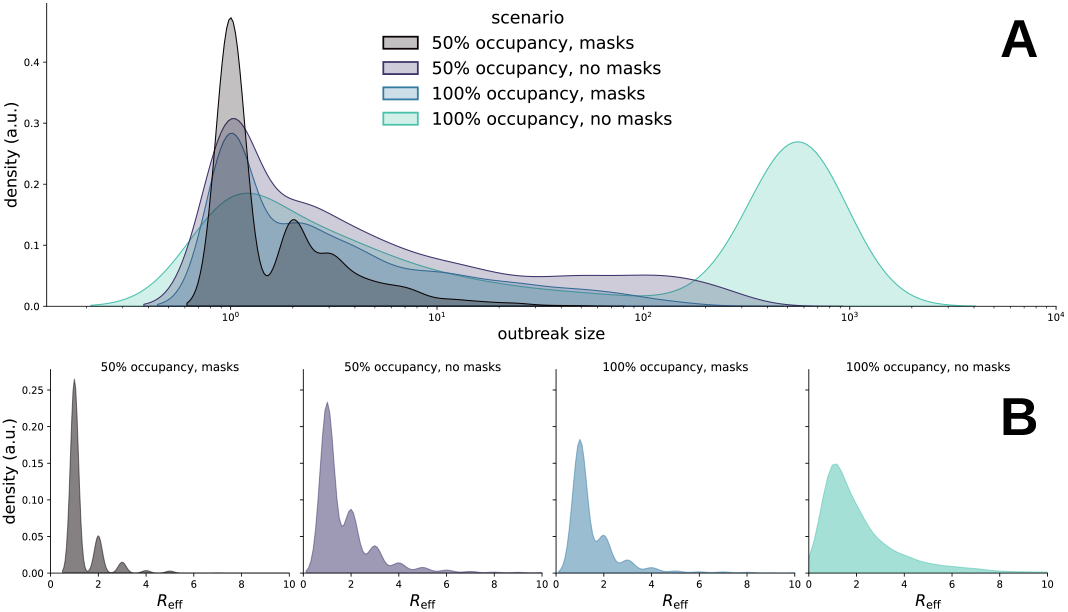
Distribution of outbreak size (**A**) and effective reproduction number *R*_eff_ (**B**) over 10 000 simulations for different lecture hall occupancy levels and masking mandates at 80% vaccination coverage for students and faculty

## Discussion

Decreasing lecture hall occupancy and imposing masking mandates are two of the most widespread policies to control the spread of COVID-19 adopted by universities. In Austria, current university policies tie lecture hall occupancy and mask mandates to the level of SARS-CoV-2 community transmission in their region. As Europe enters the cold season, community transmission is increasing or has stabilised on a high level, leading to reduced occupancy and widespread indoor mask mandates in universities.

Our findings suggest that some of these NPIs can safely be relaxed given the high vaccination rate among students: It is sufficient to either reduce lecture hall occupancy or impose mask mandates to keep *R*_eff_ *<* 1. When lowering the assumed vaccination coverage to 70%, only masks are sufficient to keep outbreak sizes in check. Nevertheless, our model is sensitive to the assumed effectiveness of vaccinations against infection (47%). If 60% effectiveness is assumed as was reported by an earlier study (12), no NPIs are necessary to maintain *R*_eff_ *<* 1. If third vaccination doses become widespread, this would allow for further relaxation of NPIs.

Since our model rests on a set of assumptions, an analysis of its shortcomings and the uncertainties associated with these assumptions is warranted. Our model does not include contact situations apart from courses on the university premises. This is a deliberate choice. It is obvious that students will meet each other in a multitude of other situations outside the university. Regulating these contacts is outside the sphere of influence of the university. Since the aim of this study is to provide information for policy decisions of universities, we do not include these contacts. On the other hand, we base our contact network on enrolment data of students to courses. Not all students that enrol in a course show up as there often is no mandatory attendance, therefore our contact network is an upper bound for the number of contacts caused by the university. We assume the vaccine efficacy against high viral load to be 47% 5 months after the second dose, which is also a conservative estimate since most students will have had their second dose during the summer in many national vaccination schemes. Lastly, we calibrated our model using empirical observations of cluster sizes in Austrian secondary schools in Autumn 2020 (13). This results in a transmission risk of 4.1% for a university contact. Here, we assume that the contact situation in secondary schools is comparable to courses at universities. We would expect contacts between secondary school students to be of higher intensity in terms of duration than contacts between university students, who have sparser schedules and class programs. Our simulation therefore poses a worst-case scenario that still draws a picture in which high vaccination rates allow to relax the implementation of NPIs.

Overall, our study assesses the two most common policies to curb the spread of SARS-CoV-2 in European universities, reduction of occupancy and mask mandates, in the context of high vaccine coverage and a dominant high transmissibility variant (delta). We find that given the currently reported vaccination rates of students of 70% and above, large outbreaks will become rare and universities can return to full on-campus operations if they introduce a single NPI: either a general mask mandate or a reduction of lecture hall occupancy to 50%.

## Data Availability

All data produced are available online at https://doi.org/10.17605/OSF.IO/UPX7R

## Data Archival

The simulation code is published as Python package https://github.com/JanaLasser/agent_based_COVID_SEIRX. The code and data used to simulate the transmission dynamics at TU Graz, as well as simulation results are available at https://github.com/JanaLasser/uni_SEIRX.

## Materials and Methods

### Co-location network

We use enrolment data on courses and exams (from now on called “events”) from the winter semester 2019/20 to construct the co-location network including students and faculty at TU Graz. The data included a total of 1623 courses and 5557 exams attended by students. For every event, a list of dates and locations for the winter semester 2019/20 was available. This resulted in a total of 24983 dates at which students and faculty met in the time between October 1 2019 and February 28 2020. If two students were enrolled in the same event, they were assumed to have a contact with each other and the responsible lecturers on every day on which a date for the corresponding event was recorded. If more than one lecturer was responsible for an event, they were also assumed to have a contact with each other. A small number of students (744) were enrolled in studies at other universities and only attended the occasional event at TU Graz. We removed this students and retained only the largest connected component of the co-location network, leaving a total of 10755 students and 974 lecturers. The final network contained 84.6% of students that were active (attended at least one lecture) and connections from 98.2% of the recorded events. See extended methods in the supporting information (SI) for details about the co-location network.

### A. Agent based model

We simulate the infection dynamics in the university using an agent-based model. The model includes two types of agents: students and lecturers. The model couples in-host viral dynamics with population dynamics. Depending on the viral load over the course of an infection, each agent is in one of five states: susceptible (S), exposed (E), infectious (I), recovered (R) or quarantined (X). In addition, after the presymptomatic phase, agents can stay asymptomatic (I1) or develop symptoms (I2). Agents remain in these states for variable time periods. Every agent has an individual exposure duration, *l*, incubation time (i.e., time until they may show symptoms), *m*, and infection duration, *n* (i.e., time from exposure until an agent ceases to be infectious). For every agent, we draw values for *l, m*, and *n* from previously reported distributions of these epidemiological parameters for the SARS-CoV-2 delta variant (see SI for details). Infections are introduced into the university through a single source case that can either be a student or a lecturer. The source case starts in the exposed state on day 0 of the simulation which corresponds to a randomly chosen day of the week. All other agents start in the susceptible state.

### Transmissions

During every interaction, an infected agent can transmit the infection to the agents they are in contact with (specified by the contact network). Transmission is modelled as a Bernoulli trial with a probability of success, *p*. This probability is modified by several NPIs and biological mechanisms *q*_*i*_, where *i* labels the measure or mechanism. Here, we consider five such mechanisms: the modification of the transmission risk due to the infection progression (*q*_1_), having or not having symptoms (*q*_2_), mask wearing of the transmitting and receiving agent (*q*_3_ and *q*_4_, respectively) and immunisation (*q*_5_). Therefore, the probability of a successful transmission is given by the base transmission risk, *β*, of a contact in the university context modified by the combined effect of these five NPIs or biological mechanisms,

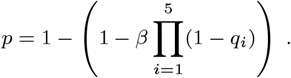

The base transmission risk *β* is calibrated to reflect the observed transmission dynamics in Austrian secondary schools (see SI for details).The calibration data was recorded in the same season as our simulation applies to (autumn), removing the need to explicitly include a seasonal effect.

### Intervention measures

In all simulations, upon first developing symptoms, agents are immediately quarantined for 10 days. There is no additional contact tracing and quarantine measures for contacts of the infected agent in place. For every event location, the seating capacity of the room is known. If occupancy is 100%, all students enrolled in a given event are allowed to attend the event, even if the number of enrolled students surpasses the seating capacity. If occupation is reduced, 50% of the students are chosen at random and their contacts are removed. At the beginning of a simulation, 80% of students and lecturers are chosen at random and assigned a “vaccinated” status. Being vaccinated reduces an agent’s chance of getting infected. Specifically, we model the effectiveness of the BNT162b2 vaccine against infection five months after the second dose of vaccination, i.e. including a drop in effectiveness with time to 47% (11). Since the viral load of vaccinated people infected with the delta variant seems to be similar to unvaccinated people (14), we do not assume a lower infectiousness of infected vaccinated agents as a conservative assumption. Vaccination status does not change throughout the simulation but vaccinated agents that are infected will recover at the end of their infection and will subsequently be completely immune to reinfection after recovery (as are unvaccinated agents that recover).

## ACKNOWLEDGMENTS

We thank Susanne Voller from TU IT Services for her help in providing the student enrolment data. We thank Harald Kainz, Rector of TU Graz, for his support of this study.

## Supplementary Information for

### Supporting Information Text

#### Extended methods

##### Co-location network

All co-location networs are available at https://github.com/JanaLasser/uni_SEIRX/tree/main/data/networks. On weekdays, students [lecturers] have an average node degree (i.e. number of contacts) of 19.8 [35.4]. As qualitatively shown in Fig. 1 of the main text, the network includes a number of communities represented by the individual degree programmes. This is also reflected in the high modularity score of 0.6, calculated with the “greedy modularity” algorithm following Clauset et al. 2004 (1)^*^. To avoid artefacts introduced by the Christmas vacation, public holidays, or reduced university operations at the beginning/end of the semester, we chose seven days from October 16 (Monday) to October 23 2019 (Sunday) for the simulation to represent a normal, full-operation week at the university. To simulate longer periods of time, we repeated this week as many times as necessary.

We note that a substantial number of students (2675) contained in the network are enrolled in a “Jointly Offered Study Programme” (“NAWI Graz”) that features lectures both at TU Graz and University of Graz. These students visit only about 50% of the lectures that are part of their degree at TU Graz and our data does not include information about the lectures provided by University of Graz. This is also reflected in their average node degree, which is only 13.7. If these students are excluded, the node degree of the remaining students is 20.7. The transmission dynamics are substantially dampened if only NAWI Graz students are considered: for example at 100% occupancy and with no masks, we find *R*_eff_ = 0.20[0; 2] and 82% of source cases do not cause an outbreak, whereas if we exclude NAWI Graz students, we find *R*_eff_ = 0.46[0; 3] and 71.9% do not cause an outbreak. Overall we note that including NAWI Graz students in the simulations yields very similar results as excluding them.

##### Calibration

We follow a slightly modified calibration approach to the version that was already described in Lasser et al. 2021 (2): As we do not include any household contacts in the contact network, we directly calibrate the transmission risk *β* for a contact in the university context. Since there is no data of sufficiently high quality about outbreaks in a higher education setting available to us, we resort to calibrating our transmission dynamics against outbreaks observed in Austrian secondary (children aged 12 to 18) and upper secondary schools (children aged 14 to 18).

To perform the calibration, we perform simulations of outbreaks in schools, relying on our modelling of the transmission dynamics in schools that has been published elsewhere (2). We compare the empirically observed and simulated distribution of cluster sizes in schools to find the best value for *β*. Specifically, we optimise the sum of the squared differences between the empirically observed cluster size distribution and the cluster size distribution from simulations *e*_1_, and the sum of squared differences between the empirically observed ratio of infected students versus infected teachers, and the simulated ratio*e*_2_. We perform simulations for values of *β* in the range of 3.81% and 5.81%, with steps of 0.17%, conducting 4000 simulations per parameter value and school type (secondary, upper secondary). We calculate the overall difference between the simulated and empirically observed cluster characteristics as

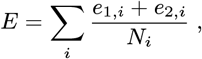

where *N*_*i*_ is the number of empirically observed clusters for school type *i*.

To assess the uncertainty associated with the optimal parameter combination we implement a bootstrapping approach on the simulated ensembles: We randomly (with replacement) sample 2,000 out of the 4,000 simulation runs for every parameter value, calculate the difference *E* between the empirically measured and simulated (subsampled) outbreak size distributions and determine the parameter value that minimises *E* for a total of 1,000 such samplings. We find a median value of *β* = 0.041[0.038; 0.046] (95% credible interval). This means that contacts in the school context have a transmission risk of 4.1% per contact.

To ensure that the thus determined optimal parameter combination is not subject to large changes if different distance measures are used to calculate *E*, we introduce additional distance measures between the empirical and simulated distributions of outbreak sizes: in addition to the sum of squared differences, we calculate the *χ*^2^ distance, the Bhattacharyya distance, the Pearson and Spearman correlation (we use the absolute value of the difference between 1 and the correlation coefficient as distance measure), and the slopes of pp- and qq-plots (we use the absolute value of the difference between 1 and the slope as distance measure). While there is some variation between the different distance measures, all confidence intervals overlap and we confirm that the optimal parameter choice only slightly depends on the chosen distance measure.

University contacts are more likely to be shorter than school contacts, since the majority of lectures, tutorials and exams last for 1-3 hours whereas students in Austrian upper secondary and secondary schools spend on average 6 hours with each other in the same classroom. The average space available for a student in a classroom of an Austrian upper secondary [secondary] school is 2.2 [2.1] m^2^, respectively, based on the average number of 23 [24] students per class (3) and the minimum room size of 50 m^2^, mandated by Austrian school building regulations (4). The average space available for a student in a lecture hall at a university (if all available seats are occupied) is 1.9 m2. Based on these facts, we assume that contacts between students, and students and students and teachers in the context of Austrian upper secondary and secondary schools are similar to contacts between students, and students and faculty in Austrian universities. If anything, we expect university contacts to be of lower intensity than school contacts due to their shorter duration.

## Footnotes

* We use the implementation of the algorithm provided by the Python networkx package: https://networkx.org/documentation/stable/reference/algorithms/generated/networkx.algorithms.community.quality.modularity.html

